# Mathematical Analysis of a COVID-19 Epidemic Model by using Data Driven Epidemiological Parameters of Diseases Spread in India

**DOI:** 10.1101/2020.04.25.20079111

**Authors:** D. Pal, D. Ghosh, P.K. Santra, G.S. Mahapatra

## Abstract

This paper attempts to describe the outbreak of Severe Acute Respiratory Syndrome Coronavirus 2 (COVID-19) via an epidemic model. This virus has dissimilar effects in different countries. The number of new active coronavirus cases is increasing gradually across the globe. India is now in the second stage of COVID-19 spreading, it will be an epidemic very quickly if proper protection is not undertaken based on the database of the transmission of the disease. This paper is using the current data of COVID-19 for the mathematical modeling and its dynamical analysis. We bring in a new representation to appraise and manage the outbreak of infectious disease COVID-19 through SEQIR pandemic model, which is based on the supposition that the infected but undetected by testing individuals are send to quarantine during the incubation period. During the incubation period if any individual be infected by COVID-19, then that confirmed infected individuals are isolated and the necessary treatments are arranged so that they cannot taint the other residents in the community. Dynamics of the SEQIR model is presented by basic reproduction number *R*_*o*_ and the comprehensive stability analysis. Numerical results are depicted through apt graphical appearances using the data of five states and India.

## 1 Introduction

Mathematical formulation of disease models is very effective to understand epidemiological prototypes of diseases, as well as it helps us to take necessary measures of public health intrusions by controlling the spread of the diseases. In early December 2019, the public of Wuhan, Hubei province in China are infected by unknown pneumonia infection ([1]-[2]). This pneumonia has been caused by novel coronavirus usually called 2019-nCoV, or SARS-CoV-2 by ICTV (severe acute respiratory syndrome coronavirus 2, by the International Committee on Taxonomy of Viruses) and it is officially named by WHO as COVID-19. From Wuhan City, COVID-19 spread rapidly whole over China [1]. As of 24:00 April 1 st, 2020 (Beijing Time), there are 81589 diagnosed cases (together with 3318 death report) in China, amid which 67,802 are from Hubei province, and over 50007 from Wuhan city, the capital of Hubei province [3]. Coronavirus was first identified in 1965, and after that three vital outbursts happened. In 2003, the first outbreak occurred in mainland China named as Severe Acute Respiratory Syndrome (SARS) ([4],[5]). The second outburst happened in 2012 at Saudi Arabia known as Middle East Respiratory Syndrome (MERS) ([6],[7]). Third outburst in the form of MERS occurred in South Korea in 2015 [8]. It was occurred fourth time in the form of COVID-19 in China and spread globally. It is noticed that the active COVID-19 patients in USA is more than the pioneering country of the outbreak China. Now this virus has started its destruction role in countries like India, Iran, Pakistan, Bangladesh, Sri Lanka and other countries of south Asia. The major symptoms of COVID-19 are almost same as SARS-CoV and MERS-CoV infections. COVID-19 patients in general suffer from dry cough, high fever, tiredness, inhalation intricacy as well as bilateral lung penetration in rigorous cases [9]. Moreover, some COVID-19 patients may suffer from nausea, vomiting as well as diarrhea without any symptoms of breathing disorder [10]. Health authority of China [11] declared that the patients primarily may give the negative result, but after subsequently tested the result may give positive result for Covid-19. After some research work ([11],[12]) it is ascertain that COVID-19 virus spreads through human to human transmission. As a result of this, the spread of this virus is not confined in China but has spread world wise. Now it has turned out to be an pandemic outburst in 205 countries with 900,306 confirmed cases as well as 45,693 confirmed deaths as of 2 April 2020 [13]. The situation is becoming worse in country like Italy, USA, Spain, France, Germany etc. Since the COVID-19 virus is transferred from human to human, the country particularly India with a high population density is in an alarming situation. If proper strategy or measures cannot be taken to protect spreading of COVID-19 virus it will be an epidemic in India and cause deaths of many Indians.

In India the first COVID-19 victim was a student from Kerala’s Thrissur district, who returned home from Wuhan University in China, and was identified as confirmed case on 30 th January, 2020 [15]. In the present senario we observe that this virus grows rapidly, moderately and slowly in some states in India. In Maharashtra (Confirmed cases-335 with deaths-16), Kerala (Confirmed cases-286 with deaths-02), Tamil Nadu (Confirmed cases-309 with deaths-01), Delhi (Confirmed cases-219 with deaths-04), Telangana(Confirmed cases-158 with deaths-07), Uttar Pradesh (Confirmed cases-172 with deaths-02), Rajasthan (Confirmed cases-167 with deaths-0) it grows very rapidly (as of 04-04-2020) [16]. It grows moderately in the states like Assam (Confirmed cases-16 with deaths-0), Bihar (Confirmed cases-29 with deaths-01),Chandigarh (Confirmed cases-18 with deaths-0), Ladakh (Confirmed cases-14 with deaths-0), West Bengal (Confirmed cases-53 with deaths-03) (as of 03-04-2020) [16]. Where as in the states like Odisha (Confirmed cases-05 with deaths-0), Puducherry (Confirmed cases-05 with deaths-0), Mizoram (Confirmed cases-01 with deaths-0), Himachal Pradesh (Confirmed cases-06 with deaths-0 1), Goa (Confirmed cases-06 with deaths-0) it grows very slowly) (as of 03-04-2020) [16].

The central government of India and all state governments have taken proper precautionary actions to restrain the spreading of COVID-19 in India from 21 March 2020. A 14-hour intended public curfew was performed in India on 22 March 2020 [17]. Also, the prime minister of India has declared complete lockdown for twenty one days starting from 24 March, 2020 [17]. The Government has take numerous procedures, like maintaining a certain social distance, encouraging social consensus on self-protection such as wearing face mask in public area, quarantining infected individuals, etc. Moreover, till the confirmed COVID-19 cases in India are growing day by day until the concluding of the current manuscript.

Now, not only medical and biological research but also mathematical modeling approach also cooperates and plays a crucial role to stop COVID-19 outburst. By mathematical modeling we may forecast the point of infection and finishing time of the disease. It also helps us to make proper decision about the necessary steps to restrain the spreading the diseases. In 2019, COVID-19 virus spreads rapidly worldwide. Therefore, it is an alarming situation for becoming a global pandemic [18] due to the severity of this virus. Therefore, real world epidemiological data is necessary for increasing situational consciousness as well as notifying involvements [19]. So far, when a pandemic such as SARS, the 2009 influenza pandemic or Ebola ([20]-[23]) outbursted, during the first few weeks of that outburst real situation analysis paid attention on the severity, transmissibility, and natural history of an budding pathogen. Again mathematical modeling supported by the dynamical equations ([24],[26]) can afford detailed characteristics of the epidemic dynamics same as statistical methods ([27]-[28]). At the early stage of COVID-19 pandemic, many researches have been done in statistical approach as well as in mathematical modeling to guess the main endemic parameters such as reproduction number, serial interval, and doubling time ([29]-[30]). Leung et al. [31] calculated number of confirmed cases transferred from Wuhan to other major cities in China. Wu et al. [32] proposed a SEIR model structure to predict disease spread world wise based on data traced from 31 December 2019 to 28 January 2020. Read et al. [25] studied a SEIR model based on COVID-19. Imai et al. [33] presented a person to person transmission COVID-19 disease and predicted the dimension of the outburst of COVID-19 in Wuhan city, China [33]. Volpert et. al. [34] studied a SIR mathematical model to restrict the increase of coronavirus via initiating firm quarantine procedures.

In this current paper, we develop a COVID-19 epidemic disease model fitted in Indian situation. In this model system we divide Indian population into five subpopulations such as susceptible population, Infected but not detected by testing population, quarantined population, Confirmed infected population who are in under treatment in isolation word, and the population who are lived in secured zone not affected by COVID-19 virus. Then by using dynamical modeling analysis, our aim is to forecast the confirmed Indian COVID-19 cases in future specifically in different States in India. We also analyze the proposed disease model mathematically to understand transmission dynamics of the COVID-19 virus amongst humans.

### 2 Model Derivation of Novel Coronavirus Disease

Several researchers have previously devised mathematical model for spread of infectious diseases ([35][38]). India is also affected by this imported disease and the number of active COVID-19 patients increases day by day right now. Depending on the recent situation, India Government has taken some strategies to stop spreading COVID-19 virus. This section presents a SEQIR model of COVID-19 based on the current situation of the disease in Indian environment. We espouse an alternate that reproduces several key epidemiological properties of COVID-19 virus. The present model structure of COVID-19 describes the dynamics of five sub-populations of Indians such as susceptible (*S*(*t*)), Infected but not detected by testing population (*E*(*t*)), quarantined (*Q*(*t*)), Confirmed infected population who are in under treatment in isolation word (*I*(*t*)) as well as population who are lived in secured zone not affected by COVID-19 virus (*R*(*t*)). We assume total population size of India is *N*(*t*) and *N*(*t*) *= S*(*t*) + *E*(*t*) + *Q*(*t*) +*I*(*t*) + *R*(*t*). In this model, quarantine refers to the separation of infected individuals from the common Indian population when the populace is infected but not infectious. By Infected Indian population, we guess that the Indian individual who have confirmed infected by the COVID-19 virus. Again by population in secured zone, we presume those Indian individuals who have not affected by corona virus disease. To make our proposed SEQIR model more realistic, we include several demographic effects by supposing a proportional natural death rate *d*_1_ > 0 in each of the five Indian sub-populations. Furthermore, we incorporate a net inflow of susceptible Indian individuals into the county (India) at a rate Λ (> 0) per unit time. Λ comprises of new birth of Indian child, immigration and emigration from and in India. The flow diagram of the COVID-19 infection model in present situation of India is depicted through Figure 1

**Fig 1:**
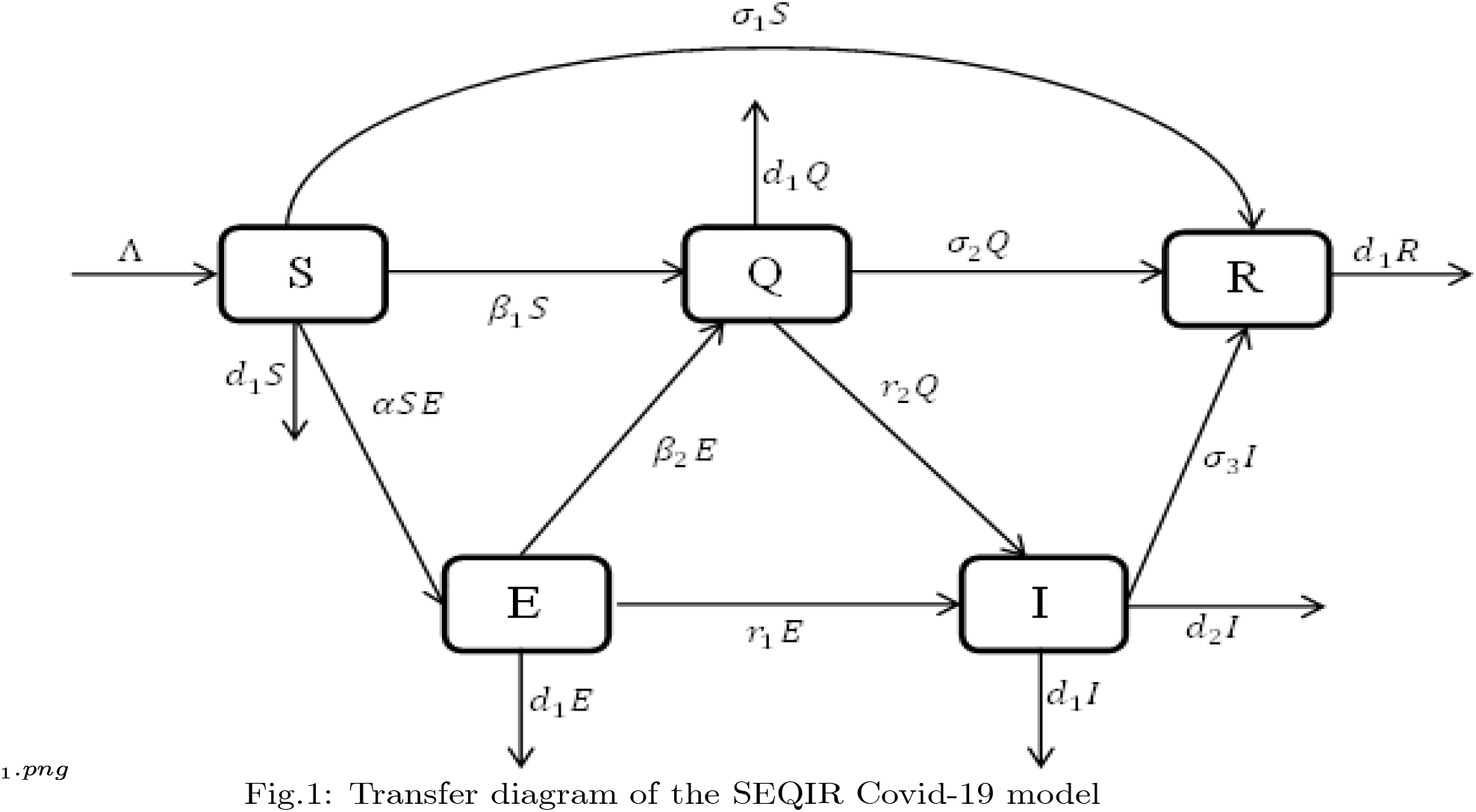
Transfer diagram of the SEQIR Covid-19 model.

#### Modeling of susceptible population (S(t))

By recruiting individuals into the region (India) at a rate Λ, the susceptible population is augmented and condensed by natural death *d*_1_. Also the susceptible population decreases through interaction between a susceptible individual and infected but not detected by testing individual. This population is also decreased by constant rates *β*_1_ and *σ*_2_ respectively to be converted into quarantined individual, as well as recovered individual. It is a real fact in India especially in the districts (Purulia, Murshidabad, Birbhum (West Bengal), Gaya, Bhagalpur (Bihar), etc.) that the susceptible population is directly sent to secured zone population due to fear effect among the inhabitants. This situation arises due to lack of proper or adequate treatment or testing facility for the large number of population in India. Therefore, the rate of change of susceptible population are governed by the following differential equation:

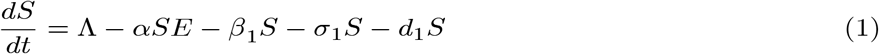

#### Modeling of Infected but not detected by testing population (E(t))

The infected but not detected by testing population indicates those individuals who are infected but their infection is not detected due to inadequate testing facility. This population increases at a rate *α* by the interaction between a susceptible individual and infected but not detected by testing individual. This population decreases due to quarantine at a rate *β*_2_ and due to natural death rate *d*_1_. Due to very high population density, it is very difficult for the Indian Government to isolate some infected but not detected by testing individual, and send them for quarantine period. Keeping this fact in mind, let this population also directly decrease by infected population at a rate *r*_1_. Therefore, the rate of change of infected but not detected by testing population is governed by the following differential equation:

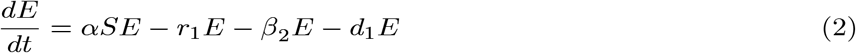

#### Modeling of quarantine population (Q(t))

Incubation period for COVID-19 is 2 days to 14 days. This period is very crucial for disease transformation from one individual to another individual. Therefore, we have to isolate those individual from susceptible and infected but not detected by testing individual for 14 days to control spread of COVID-19 in India. This mentioned population is known as quarantined population. Quarantined population is increased at a rate *β*_1_ and *β*_2_ from susceptible as well as infected but not detected by testing population respectively. This population is decreased at a rate *r*_2_ and *σ*_2_ due to infected population and population in secured zone correspondingly. Let the natural death rate be *d*_1_ of this population, hence the rate of change of quarantine population is as follows:

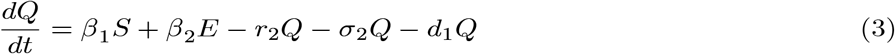

#### Modeling of confirmed infected population (I(t))

The infected population who have confirmed positive report by the COVID-19 test, is increased by infected but not detected by testing at a rate *r*_1_ (infected but not detected by testing, such all population are not possible for quarantined due to lack of space or other reasons. Infected but not detected by testing individual may become illness for COVID-19, and subsequently their test report becomes positive. So they enter directly to the infected population, this particular case is very harmful to protect spreading COVID-19 in India), and also increased at rate *r*_2_ from quarantined population as usual. Infected population is decreased at rate *σ*_3_. and *d*_1_ due to recovered population respectively. Let the natural death rate be *d*_1_ and to make it more realistic *d*_2_ is the rate of death for infection, and hence the rate of change of infected population is governed by the following differential equation:

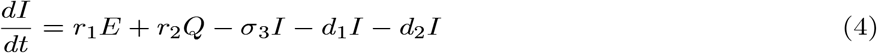

#### Modeling of secured zone population not affected by COVID-19 (R(t))

We assume that susceptible, quarantine as well as infected individuals recover from the disease at rates *σ*_1_,*σ*_2_ and *σ*_3_ respectively and enter in secured zone population. This population is reduced by a natural death rate *d*_1_. Thus, rate of change of secured zone population is not affected by COVID-19 virus is governed by the following differential equation:

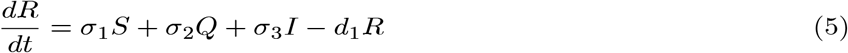

Combining equations ((1)-(5)) our wished-for model structure takes the following form:

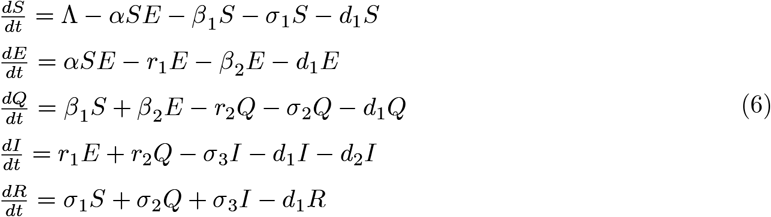

with initial densities:

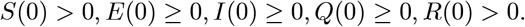

All the parameters and corresponding biological meaning are presented in Table 1 given below.

**Table 1:** Explanation of parameters exploited in our proposed SEQIR model structure.

|  |  |
| --- | --- |
| Parameters | Meaning |
| $\Lambda$ | The recruitment rate at which new individuals enter in the Indian population |
| $\alpha$ | The transmission rate from susceptible population to infected but not detected by testing population |
| $\beta_1$ | The transmission coefficient from susceptible population to quarantine population |
| $\beta_2$ | The transmission coefficient from infected but not detected by testing population to quarantine population |
| $\sigma_1$ | The transmission rate from susceptible population to secured zone population |
| $\sigma_2$ | The transmission coefficient from infected but not detected by testing population to secured zone population |
| $\sigma_3$ | The transmission rate from quarantine population to secured zone population |
| $r_1$ | The transmission rate from infected but not detected by testing population to infected population for treatment |
| $r_2$ | The transmission rate from quarantine population to infected population for treatment |
| $d_2$ | Death rate of Infected population due to Covid-19 infection |
| $d_1$ | Natural death rate of all five sub-populations |

The above SEQIR model formulation (6) can be rewritten as

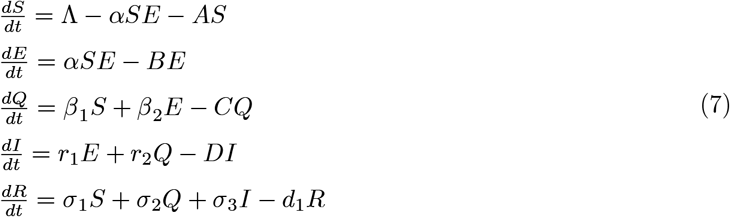

where *A* = (*β*_1_ + *σ*_2_ + *d*_1_), *B* = (*r*_1_ + *β*_2_ + *d*_1_), *C =* (*r*_2_ + *σ*_2_ + *d*_1_) and *D* = (σ_3_ + *d*_1_ + *d*_2_).

## 3. Basic Properties

### 3.1 Non negativity of solution

#### Theorem 1

*Each solution of the SEQIR model structure* (*7*) *with preliminary stipulations subsists in the interval* [0, ∞) *and* S(*t*) > 0, *E*(*t*) ≥ 0, *I*(*t*) ≥ 0 *as well as R*(*t*) > 0 *for all values *t* greater than equal to zero*.

**Proof**. As the right hand side of SEQIR model structure (7) is completely continuous and locally Lipschitzian on *C*, the solution (*S*(*t*),*E*(*t*),*I*(*t*),*R*(*t*)) of (7) with initial conditions exists, and is unique on [0, *ξ*), where 0 < *ξ* < +∞.

From system (7) with initial condition, we have

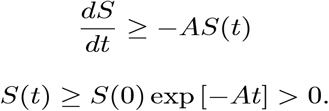

The second equation of system (7), implies 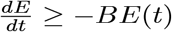

Using initial condition we obtain *E*(*t*) ≥ *E*(0) exp [−*Bt*] ≥ 0

Again from third and fourth equations of the system (7) with the help of initial condition provide

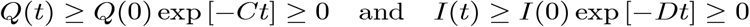

Furthermore, last equation of (7) provides 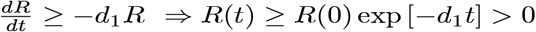

Therefore, we can see that *S*(*t*) > 0, *E*(*t*) ≥ 0,*I*(*t*) ≥ 0, *R*(*t*) > 0, ∀*t* ≥ 0. This concludes the proof of the theorem.

■

### 3.2 Invariant region

#### Theorem 2

*All solutions of the SEQIR model structure* (*7*) *that initiate in* 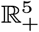 *are bounded and enter into a region* Ω *defined by* 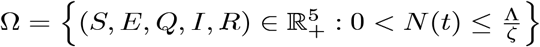 *t* → ∞, *where ζ* = min {*d*_1_, *d*_1_ + *d*_2_].

**Proof**. Suppose *N*(*t*) = *S*(*t*) + *E*(*t*) + *I*(*t*) + *R*(*t*), where (*S*(*t*),*E*(*t*),*Q* (*t*), *I*(*t*),*R*(*t*)) SEQIR model structure (7). Differentiating both sides with respect to *t*, we have

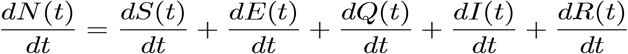

After substitute the values of 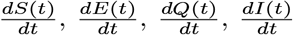 and 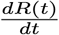 form equation (7), we obtain

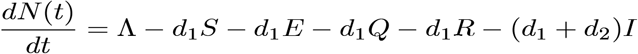

Now

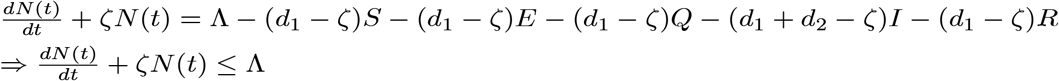

where *ζ* = min {*d*_1_, *d*_1_ + *d* _2_}. Now by using comparison theorem, we obtain

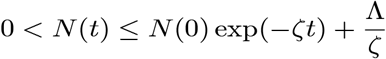

As *t* → ∞, we have 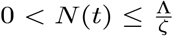. Therefore, all solution of the model structure (7) enter in the region 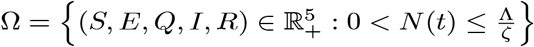 Hence, we have completed the required proof.

■

## 4 Diseases-Free Equilibrium and the Basic Reproduction Number

The diseases-free equilibrium (DFE) of the proposed SEQIR model (7) is acquired by setting *E* = 0, *Q* = 0, I = 0 and *R* = 0. The obtained DFE is 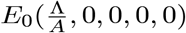.

Basic Reproduction Number (BRN) ([39]-[42]) is a number which is defined as the new infective formed by a solitary infective individual for the duration of his or her effectual infectious epoch when introduced into an utterly susceptible populace at equilibrium.

To find the BRN of our proposed SEQIR model structure (7), we take the assistance of next-generation matrix method [41] formulation. Assume *y* = (*E, Q, I, R, S*)^*T*^, then the system (7) can be rewritten as

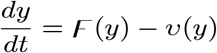

where 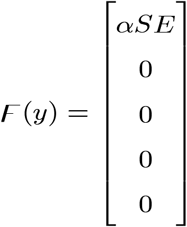 and 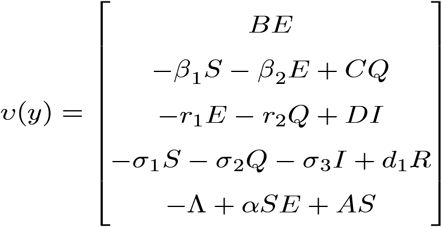

ℱ is known as transmission part which, articulates the production of new infection and *v* is known as transition part, which explain the alter in state.

The Jacobian matrices of ℱ (*y*) and *v*(*y*) at the DFE *E*_0_ is given by

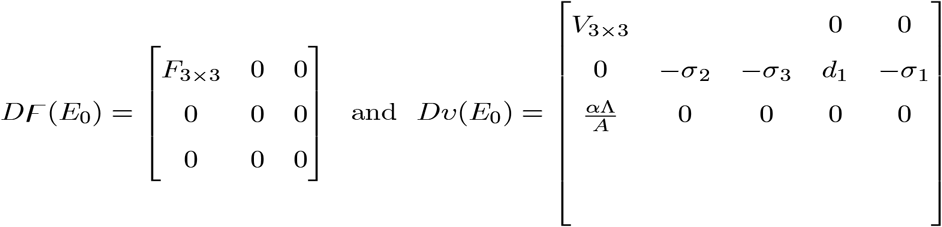

where 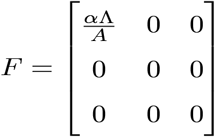 and 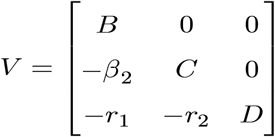

Therefore, *FV* ^−1^ is the next generation matrix of the SEQIR model structure (7). So, as per [41] *R*_0_ = *ρ* (*FV* ^−1^) where *ρ* stands for spectral radius of the next-generation matrix *FV* ^−1^. Therefore,

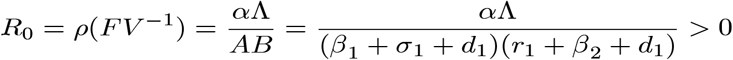

It is notable that 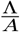 represents the number of susceptible individual at the DFE *E*_0_.

### 4.1 Sensitivity Analysis of *R*_0_

Based on the each of the parameters of *R*_0_ a sensitivity analysis is performed to check the sensitivity of the basic reproduction number. As indicated by Arriola and Hyman [43] we have computed the normalized forward sensitivity index with respect to each of the parameters.

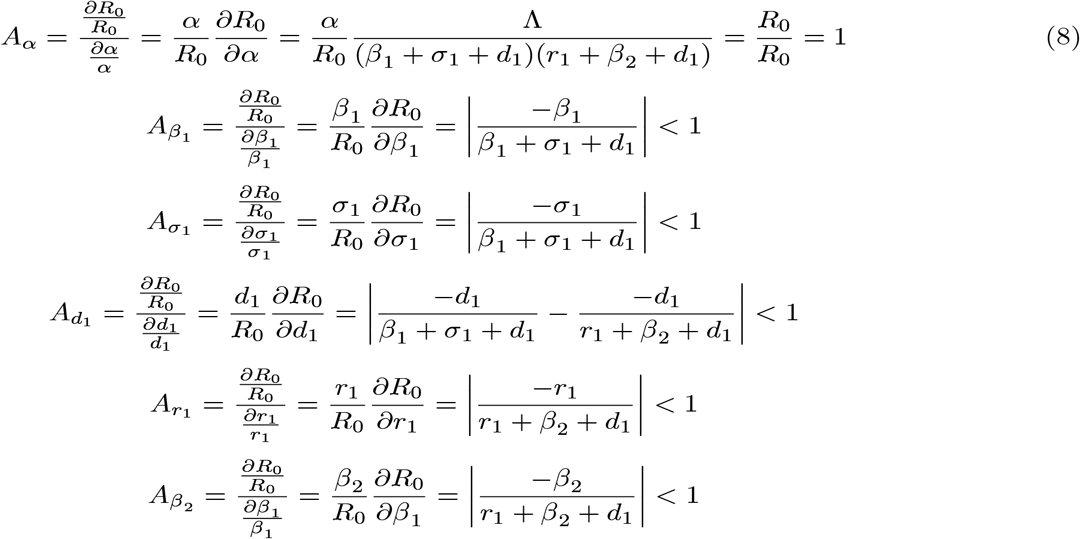

From the above calculations, it is obvious that the BRN *R*_0_ is mainly sensitive to alters in *α*. The value of *R*_0_ will be enhanced if we raise the value of *α*. On the contrary, the value of *R*_0_ will be as well reduced in the same proportion if we diminish the value of *α*. Again it is also noticed that the parameters *β*_1_, *σ*_1_, *d*_1_, *r*_1_ and *β*_2_ are related to *R*_0_ inversely. Therefore, for any increasing value of any of this mentioned parameters will definitely reduce the value of *R*_0_. But the decreasing value of *R*_0_ will be comparatively slighter. Since the effect of parameters *β*_1_, *σ*_1_, *d*_1_, *r*_1_ and *β*_2_ are very small on *R*_0_, it will be sensible to focus efforts on the reduction of *α* (transmission rate at which the susceptible individual converted to exposed individual). Therefore, the sensitive analysis of the basic reproduction number emphasized that prevention is better than treatment. That is exertions to enhance prevention are further effective in controlling the spread of COVID-19 disease than to enlarge the numbers of individuals accessing treatment, as yet there is no proper vaccine, which is medically proven this technique is very much effective to control the spread of Coronavirus disease in country like India. Following the same concept, the Indian Government as well as all State Government of India have taken necessary action such as Lockdown, campaign against the disease etc. to protect the out break of this dangerous disease.

### 4.2 Stability Analysis of Diseases-free Equilibrium

This section is constructed with an aim to study the stability nature of the Diseases Free Equilibrium (DFE) *E*_0_ with the assistance of subsequent Theorems:

#### Theorem 3

*The SEQIR model structure* (*7*) *at* 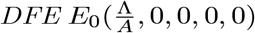 *is locally asymptotically stable under the condition R*_0_ < 1 *and became unstable if R*_0_ > 1.

***Proof***. *The Jacobian of the system* (*7*) *at* 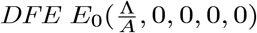 *is given by*

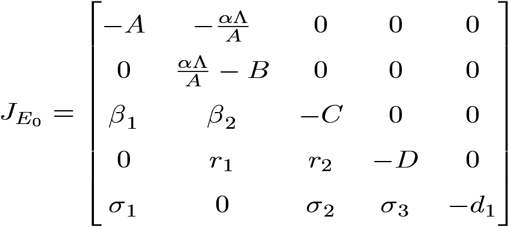

*The characteristic equation of the matrix* 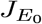 *is given*

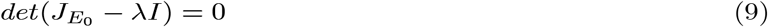

*where λ is a eigenvalue of the matrix* 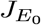. *Therefore, roots of the equation* (*9*) *i*.*e, eigenvalues of the matrix* 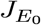 *are* 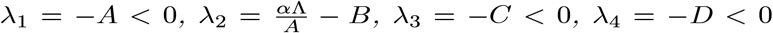 *as well as λ*_5_ = − *d*_*i*_ < 0.

*Therefore, Therefore the system* (*7*) *is locally asymptotically at the* 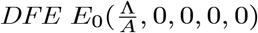. *Now λ*_2_ < *0*

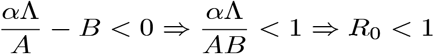

*Hence*, 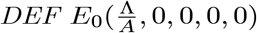 *is locally asymptotically stable under the condition R*_0_ < 1.

■

#### Theorem 4

*The SEQIR model structure* (*7*) *at* 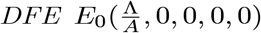 *is globally asymptotically stable* (*GAS*) *under the condition R*_0_ < 1 *and became unstable if R*_0_ > 1.

**Proof**. We can rewrite the system of differential equation (7) in the following form given below

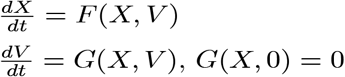

Here, *X* = (*S, R*) ∈ *R*^2^(the number of uninfected individuals compartments), *V* = (*E, Q, R*) ∈ *R*^3^(the number of infected individuals compartments) as well as 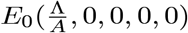 is the DFE of the SEQIR model structure (7). DFE *E*_0_ is globally stable if the subsequent two stipulations are fulfilled:

1. 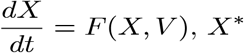 is globally asymptotically stable,
2. 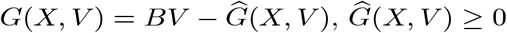,

where *B* = *D*_*V*_*G*(*X*^∗^, 0), is a Metzler matrix and Ω is the positively invariant set with respect to the model (7). Following Castillo-Chavez et al. [44], we check for aforementioned conditions. For our proposed model structure(7) 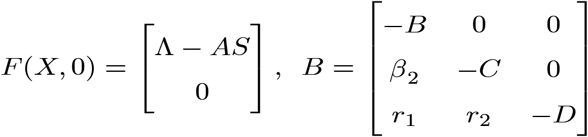 and 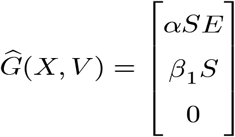

Obviously, 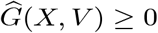 every time the state variables are within Ω. It is also obvious that 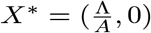 is a globally asymptotically stable equilibrium of the system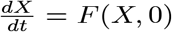. Hence, the theorem is proved.

## 5 Existence of Endemic Equilibrium Point and its Stability

This section reflects the existence of endemic equilibrium as well as demonstrates its stability nature. To stumble on the endemic equilibrium *E*_1_(*S*^∗^, *E* ^∗^, *Q*^∗^, *I* ^∗^, *R*^∗^) of our wished-for SEQIR structure (7) we deem the following:

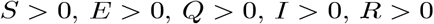

as well as

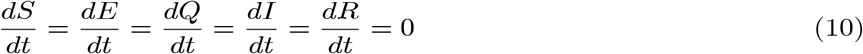

Solving the system of equation (10) we obtain

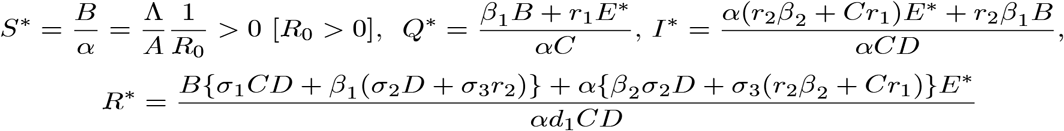

From first equation of (10) we have

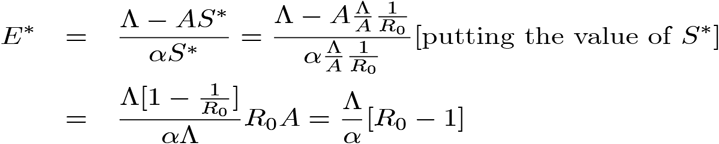

Therefore, *E*^∗^ has unique positive value if *R*_0_ − 1 > 0 i.e., *R*_0_ > 1. Shortening the above discussions and discussions in the previous sections we have reached the following theorem:

### Theorem 5

*The SEQIR model structure* (*7*) *has a unique* 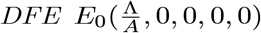 *for all parameter values as well as the system* (*7*) *has also a unique endemic equilibrium E*_*X*_(*S*^∗^, *E* ^∗^, *Q*^∗^, *I* ^∗^, *R*^∗^) *under the condition R*_0_ > 1.

The next theorem establish the local stability nature of the endemic equilibrium *E*_1_ (*S*^∗^, *E*^∗^, *Q*^∗^, *I* ^∗^, *R*^∗^)

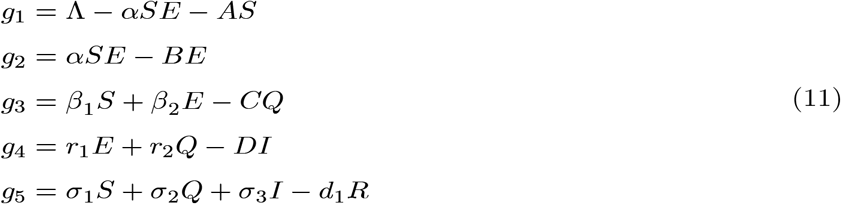

We assume *α*^∗^ = *α* as the bifurcation parameter, predominantly as it has been explained in (8) that *R*_0_ is extra sensitive to alter in than the other parameters. If it is considered that *R*_0_ = 1, then we have

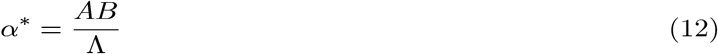

Now, the Jacobian of the linearized system (11) via (12) at DFE *E*_0_ when *α*^∗^ = *α*, is as follows:

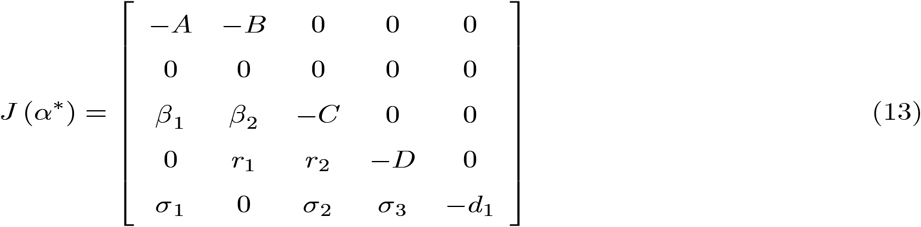

Eigenvalues of the matrix (13) are given by (0, − *A*, − *C*, − *D*, − *d*_1_)^*T*^. We observe that the matrix (13) has simple zero eigenvalue, and the other eigenvalues are negative. We are now at the stage to apply center manifold theory [45] to analyze the dynamics of system (11). Corresponding to zero eigenvalue the right eigenvector *ω* = (*ω*_1_, *ω*_2_, *ω*_3_, *ω*_4_, *ω*_5_)^*T*^ of the matrix (13) is given by

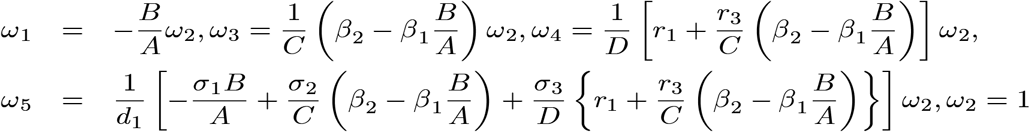

with *ω*_2_ free. Moreover, *J* (*α*^∗^) has a corresponding left eigen vector *v =* (*v*_1_, *v*_2_, *v*_3_, *v*_4_, *v*_5_) where *v*_1_ = 0, *v*_2_ = 0, *v*_3_ = 0, *v*_4_ = 0, *v*_5_ =0

with *v*_2_ free. Therefore, we have

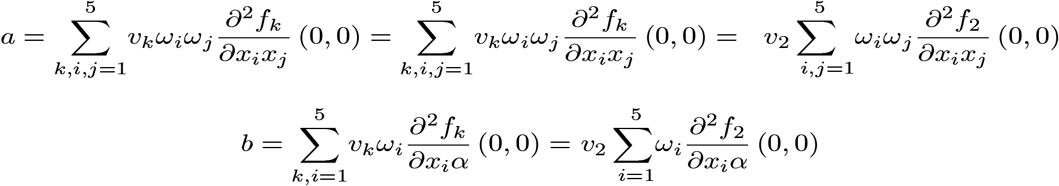

Substituting the values of all the second-order derivatives calculated at DFE as well as *α*^∗^ = *α* we obtain,

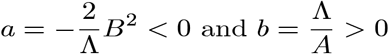

As *a* < 0 and *b* > 0 at *α*^∗^ = *α*, hence, from Remark 1 of the Theorem 4.1 as declared in [44], a transcritical bifurcation takes place at *R*_*o*_ = 1 as well as the inimitable endemic equilibrium is locally asymptotically stable due to *R*_*o*_ > 1. Hence, we conclude the proof.

## 6 Numerical Verification and Predictions

To fit our proposed COVID-19 model system (6) to the daily new COVID cases for all over India as well as five states Delhi, Kerala, Maharashtra, Uttar Pradesh and West Bengal, we performed numerical simulations in this section. Data are collected from the official website of Indian Council of Medical research and World Health Organization ([47],[48]). We first estimated the values of different parameters of the model given in table 2.

**Table 2:**
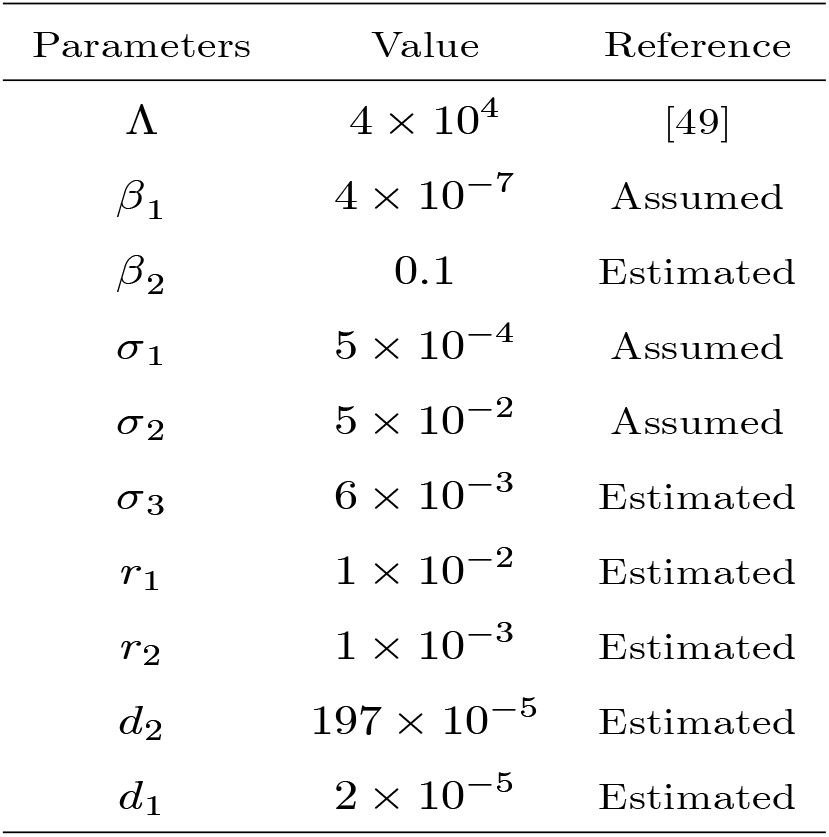
Values of the parameter of the model system (6) for India.

| Parameters | Value | Reference |
| --- | --- | --- |
| $\Lambda$ | $4 \times 10^4$ | [49] |
| $\beta_1$ | $4 \times 10^{-7}$ | Assumed |
| $\beta_2$ | 0.1 | Estimated |
| $\sigma_1$ | $5 \times 10^{-4}$ | Assumed |
| $\sigma_2$ | $5 \times 10^{-2}$ | Assumed |
| $\sigma_3$ | $6 \times 10^{-3}$ | Estimated |
| $r_1$ | $1 \times 10^{-2}$ | Estimated |
| $r_2$ | $1 \times 10^{-3}$ | Estimated |
| $d_2$ | $197 \times 10^{-5}$ | Estimated |
| $d_1$ | $2 \times 10^{-5}$ | Estimated |

Initial densities as of 21th March 2020 is given in Table-3

In our proposed model system (6) most sensitive parameter is *α* (transmission rate from susceptible population to infected but not detected by testing population). Therefore, we mainly aspire to see the effect of on COVID-19 disease spreading. For different values of *α*, the value of *R*_0_ is presented in Table 4

**Table 3:** Initial densities of the model system (2.6) for India.

| $S(0)$ | $E(0)$ | $Q(0)$ | $I(0)$ | $R(0)$ |
| --- | --- | --- | --- | --- |
| $8 \times 10^8$ | $15 \times 10^2$ | $5 \times 10^4$ | 284 | $4 \times 10^8$ |

**Table 4:** Value of *R*_0_ for different *α* for India.

| $\alpha$ | $2.1 \times 10^{-10}$ | $2.5 \times 10^{-10}$ | $2.8 \times 10^{-10}$ |
| --- | --- | --- | --- |
| $R_0$ | 0.35847 | 0.42675 | 0.47796 |

From the above Table we observe that all values of *R*_0_ is less than unity. Hence, the endemic equilibrium *E*_1_ is not locally asymptotically stable for these values of *R*_0_ and corresponding *α*. Now we plot the graph of *R*_0_ with respect to *α*.

From the above figure we observe that as the values of *α* increases *R*_0_. It is also observed that after certain value of *α, R*_0_ becomes greater than 1. Therefore, up to certain value of *α* DFE is stable (Theorem 3), and beyond that value of *α* DFE becomes unstable. In this current situation of the universe, we are interested about the infected population *I*(*t*) as days progress. Therefore, drawing the time series plot of infected population taking initial densities as given in Table 3.

The figure 3 is very interesting because we see that for *α =* 2.5 × 10^−l0^ the confirmed infected population of our proposed model system exactly fitted to the real confirmed infected individuals in India so far. To show the peak of the infection we have drawn the Figure 4 for *α =* 2.5 × 10^−l0^.

**Figure 2:**
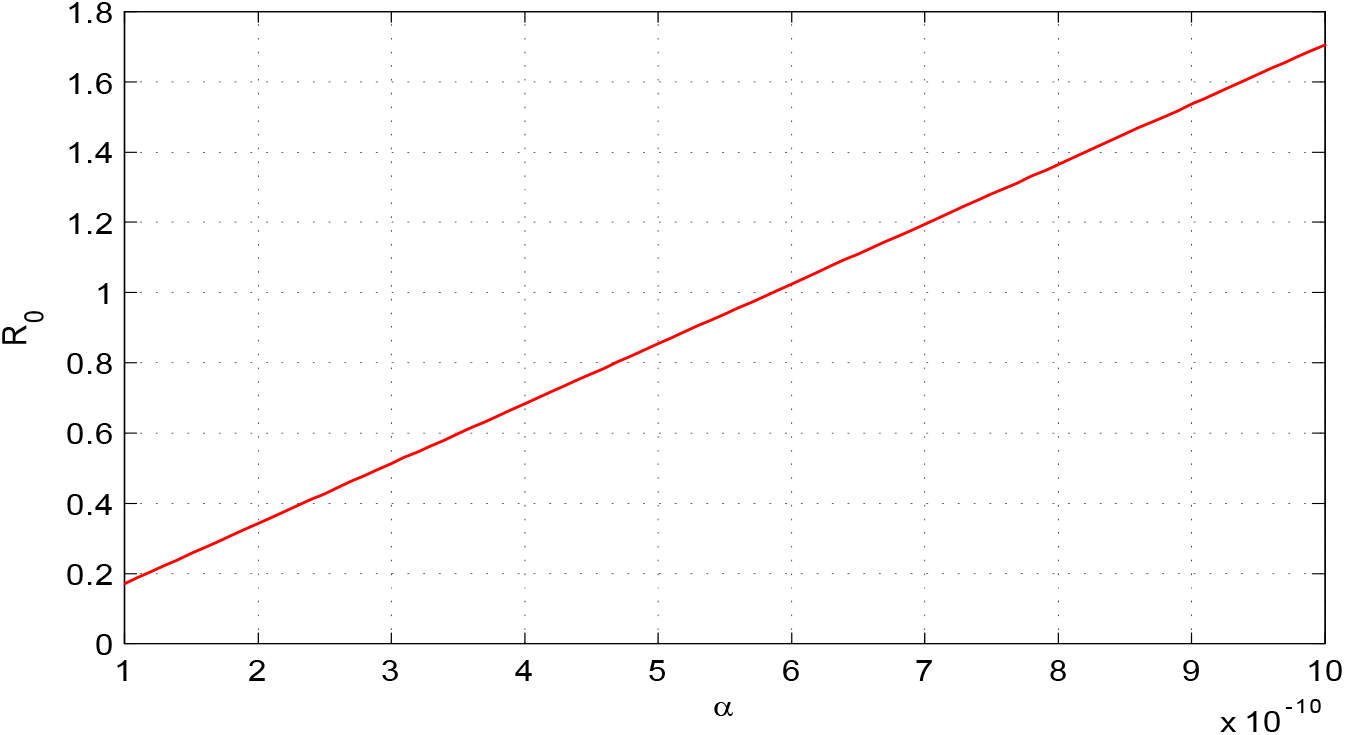
Variation of *R*_0_ with respect to most sensitive parameter *α*.

**Figure 3:**
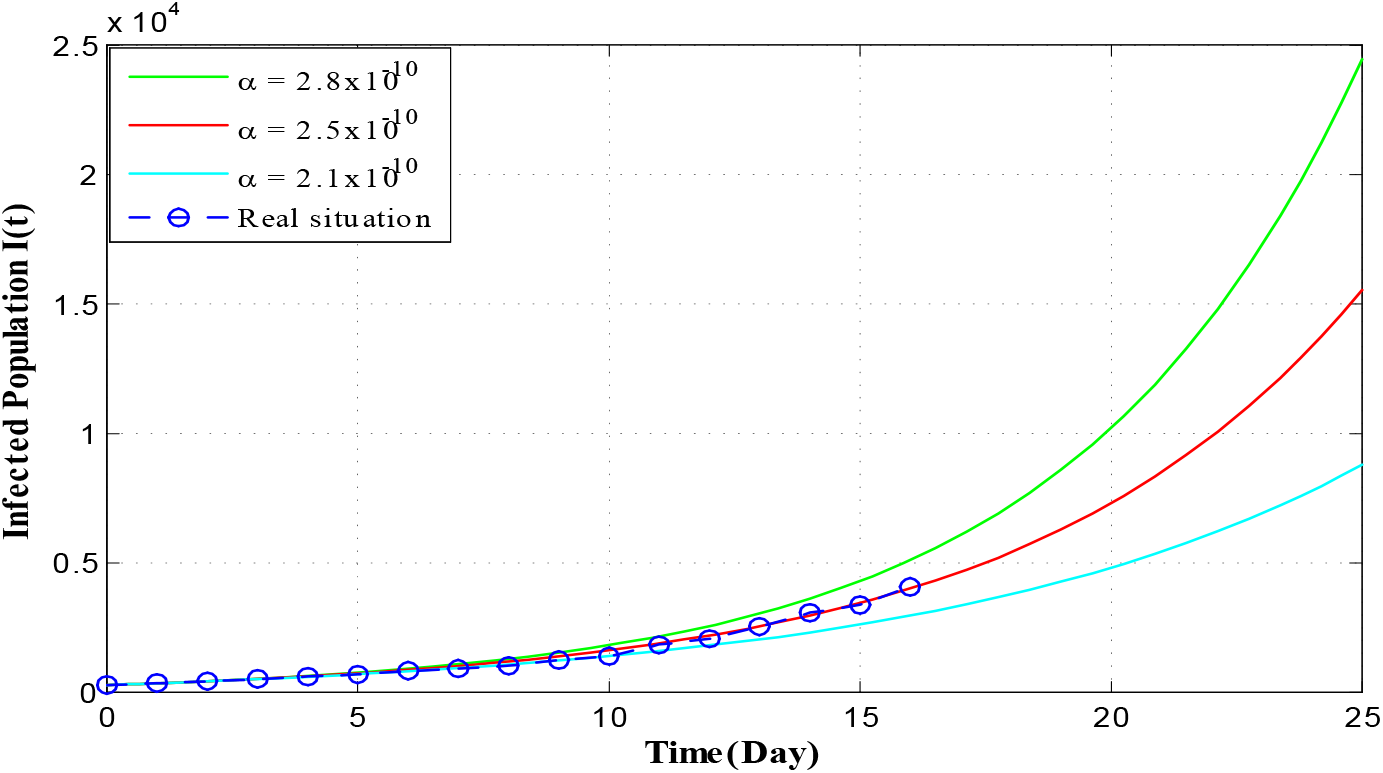
Time series plot of infected population for *α* + 2.8 × 10^−l0^, *α* + 2.5 × 10^−l0^ and *α* + 2.1 × 10^−l0^ with initial conditions, parameter values are given in Table 2 and Table 3 respectively.

Figure 4 of the infected population of our proposed model shows that, imposing the restricting measures from 21 March, 2020 by the Government of India, the peak of infection (maximum of daily cases) is attained almost after four months (120 days). After that if same restriction is continued, the peak of infection is gradually decreases.

**Figure 4:**
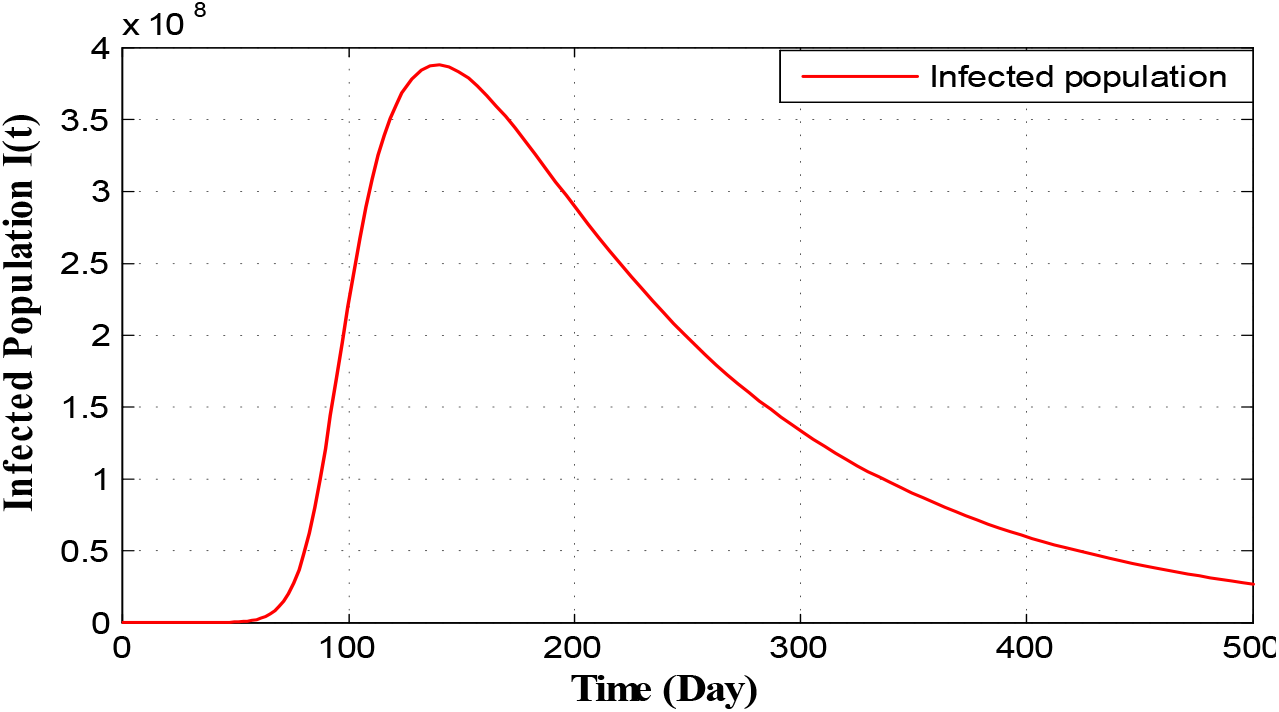
Long run time series plot of infected population for *α* + 2.5 × 10^−l0^ with initial conditions, parameter values are given in Table 2 and Table 3 respectively

The short time behavior of our proposed model i.e., after 30 days and 60 days from the restriction measure started by the Indian Government, estimated infected individuals are depicted through Figure 5 and Figure 6 respectively.

**Figure 5:**
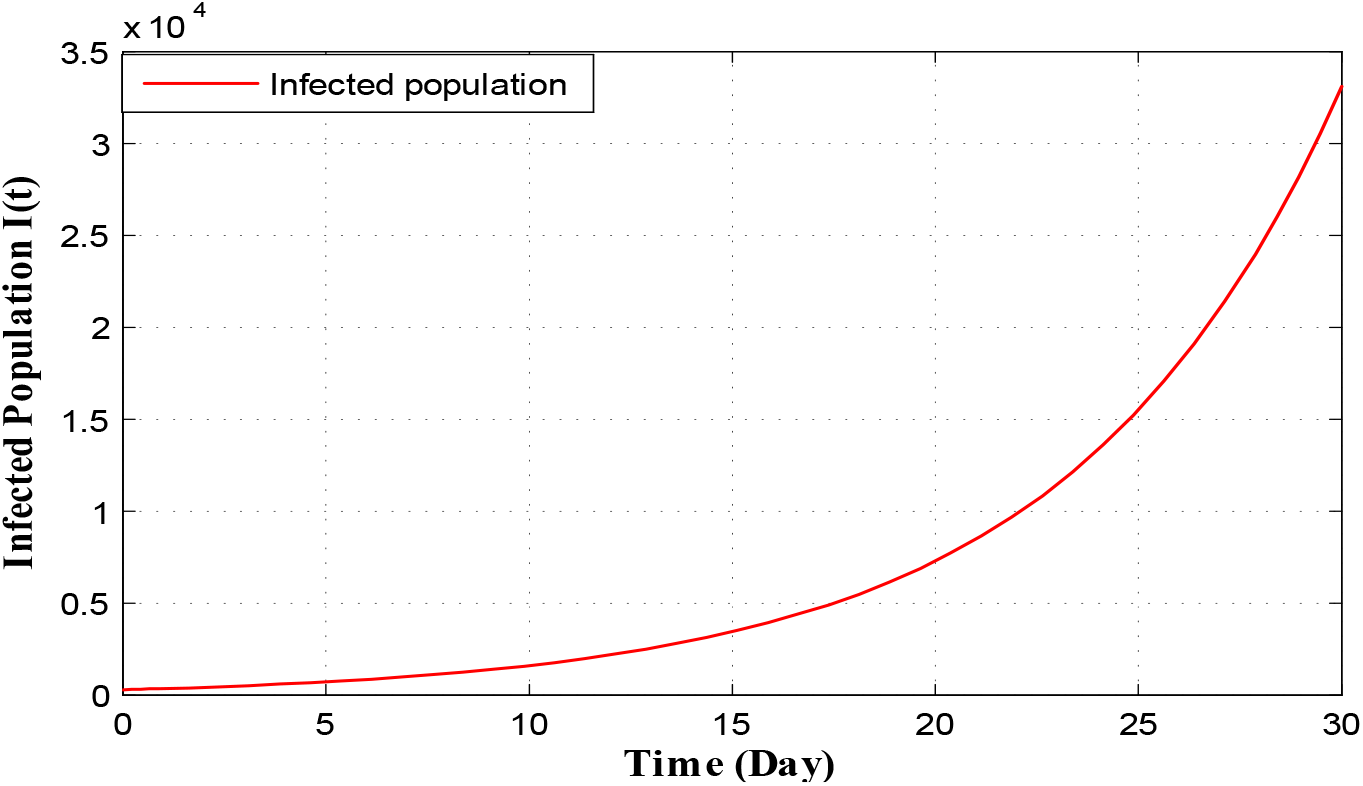
Time series plot of infected population for *α* + 2.5 × 10^−l0^ with initial conditions, parameter values are given in Table 2 and Table 3 respectively for one month period

**Figure 6:**
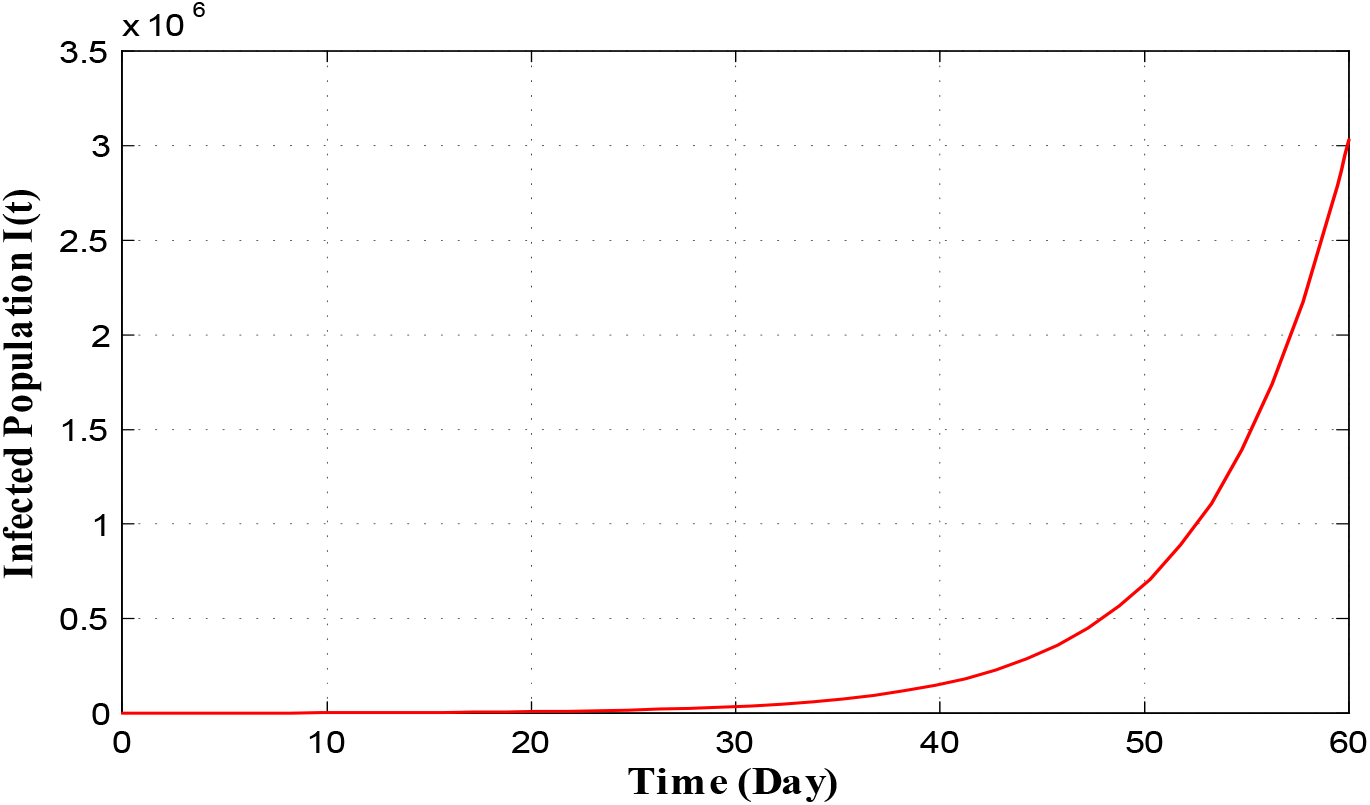
Time series plot of infected population for *α* + 2.5 × 10^−l0^ with initial conditions, parameter values are given in Table 2 and Table 3 respectively for two months period

Therefore, implementing Government restrictions mathematically, Fig 5 and Fig 6 infers that in thirty days number of infected individual in India goes to about 32000. In 60 days this number goes to 3000000. Since, Government is changing its restricting measures after certain period of time policies, so the system parameters are also altering after that period of time. Therefore, in the current position we are not interested about infected population for long time of period.

Now, we simulate the daily new confirmed COVID-19 cases for the five states of India namely Maharashtra, Kerala, Uttar Pradesh, Delhi and West Bengal starting from 21 March, 2020. We try to fit the model (6) to day by day novel confirmed COVID-19 cases in the five states of India. Here we also taken *α* (transmission rate from susceptible population to infected but not detected by testing population) as before. Estimated initial densities as of 21 March, 2020 are given in Table 5. Estimated Recruitment rates and infection tempted mortality rates for the five states of India are given in Table 6. Lastly, rests of the estimated parameters are given in Table 7.

**Table 5:** Estimated initial densities (as of 21 March, 2020)

| State | $S(0)$ | $E(0)$ | $Q(0)$ | $I(0)$ | $R(0)$ |
| --- | --- | --- | --- | --- | --- |
| Maharashtra | 75000000 | 225 | 800 | 58 | 30000000 |
| Kerala | 10000000 | 200 | 1000 | 40 | 10050000 |
| Uttar Pradesh | 150000000 | 120 | 1500 | 24 | 50000000 |
| Delhi | 10000000 | 150 | 800 | 27 | 500000 |
| West Bengal | 50000000 | 20 | 200 | 03 | 30000000 |

**Table 6:** Recruitment rates and infection tempted mortality rates for the five states of India.

| Parameters | Maharashtra | Kerala | Uttar Pradesh | Delhi | West Bengal |
| --- | --- | --- | --- | --- | --- |
| $\Lambda$ | 3300 | 405 | 14200 | 650 | 3490 |
| $d_2$ | 0.0032 | 0.00029 | 0.0049 | 0.00067 | 0.0024 |

**Table 7:** Estimated parameters for the five states of India.

| Parameters | Maharashtra | Kerala | Uttar Pradesh | Delhi | West Bengal |
| --- | --- | --- | --- | --- | --- |
| $\beta_1$ | 0.0000004 | 0.0000004 | 0.0000004 | 0.0000004 | 0.0000004 |
| $\beta_2$ | 0.005 | 0.005 | 0.005 | 0.005 | 0.005 |
| $\sigma_1$ | 0.0005 | 0.0005 | 0.0005 | 0.0005 | 0.0005 |
| $\sigma_2$ | 0.1 | 0.1 | 0.1 | 0.1 | 0.1 |
| $\sigma_3$ | 0.005 | 0.012 | 0.0038 | 0.0016 | 0.0078 |
| $r_1$ | 0.04 | 0.05 | 0.02 | 0.05 | 0.05 |
| $r_2$ | 0.002 | 0.001 | 0.0012 | 0.00093 | 0.0005 |
| $d_1$ | 0.000015 | 0.000018 | 0.00002 | 0.00001 | 0.000016 |

Using these estimated parameters, the fixed parameters and initial densities (Table 5 to Table 7) we graphically present infected population for different states. Figure 7 and Figure 8 present the graphical presentation of infected population for the state Maharashtra

**Figure 7:**
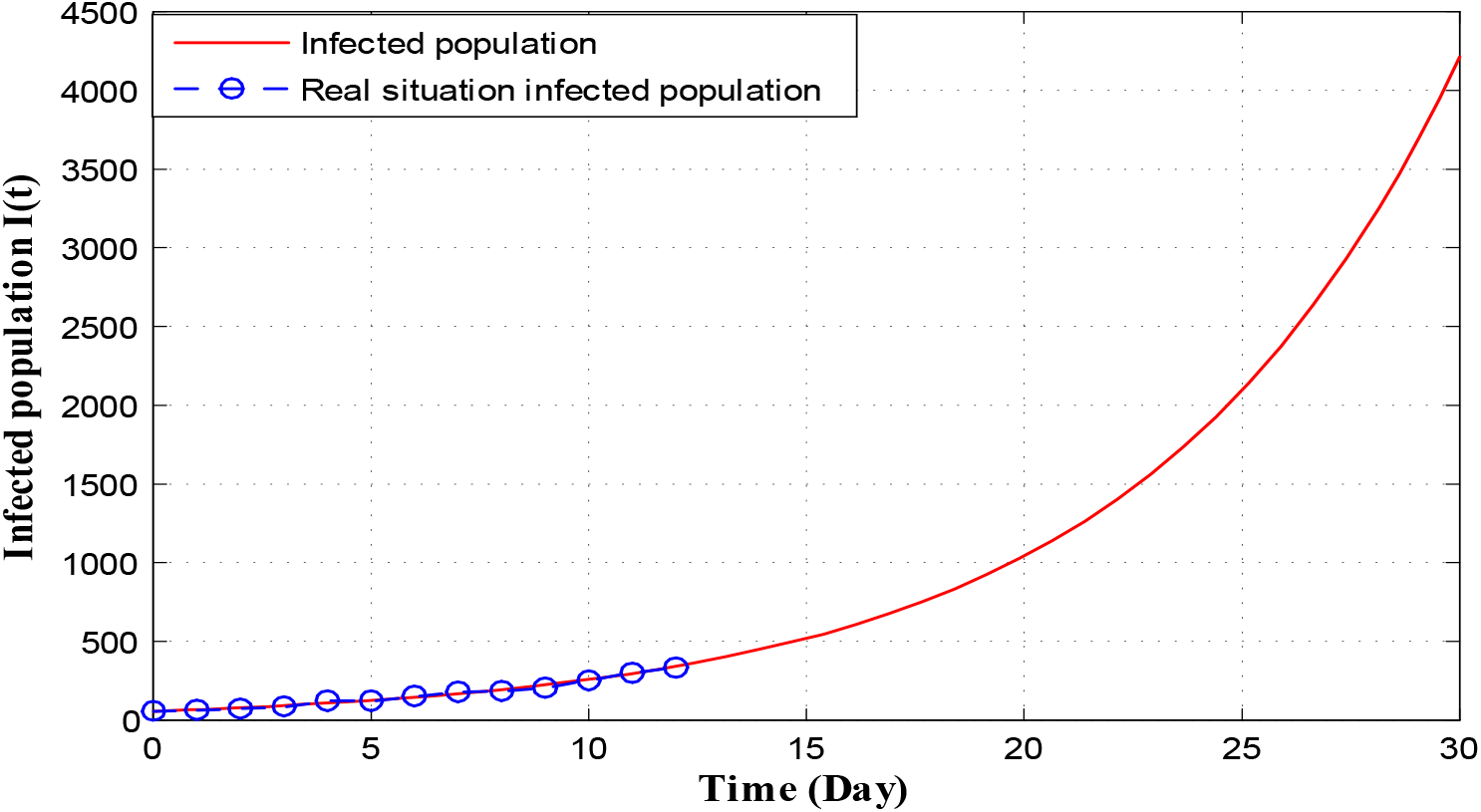
Time series plot of infected population in Maharashtra for *α =* 2.5 × 10^−9^ using data from Table 5 Table 6 and Table 7 for one month period from 22 nd March, 2020.

**Figure 8:**
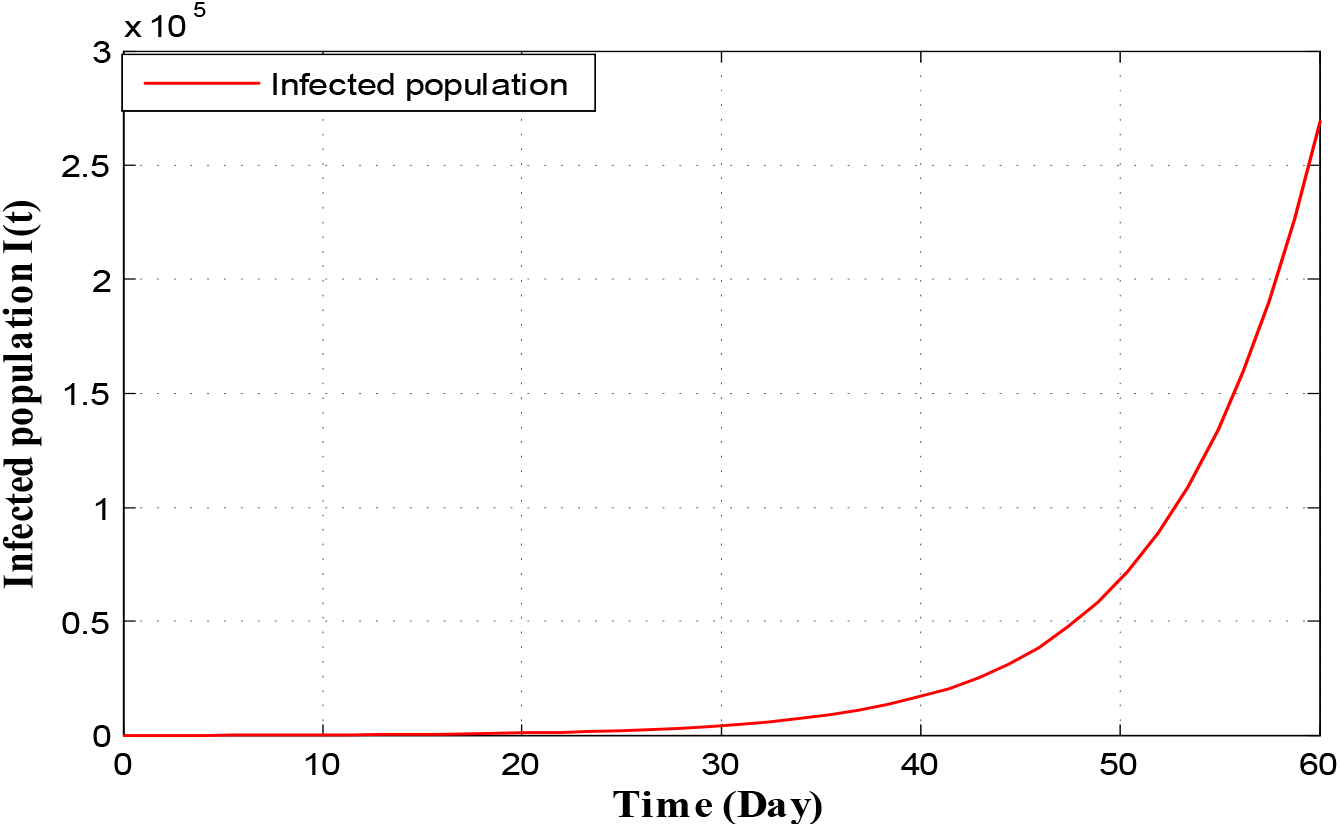
Time series plot of infected population in Maharashtra for *α =* 2.5 × 10^−9^ using data from Table 5 Table 6 and Table 7 for two months period from 22nd March, 2020

From Figure 7, we monitor that as per our imposed restricting measures as given by Government of India, total number of infected individuals in Maharashtra reaches to 4100 in thirty days from 21 March, 2020. We also detect from Figure 7 that the actual number of infected persons also coincides with the infected COVID-19 individuals of our proposed system for 13 days starting from 21 March, 2020. Again Figure 8 infers that total number of infected population reaches to 260000 in two months. But in reality this situation may not happen as Government is continuously changing its restricting measures to stop the spreading of COVID-19.

Now to illustrate the COVID-19 situation in Kerala based on values of parameters as given in Table 5 to Table 7 we draw the following two figures

The Figure 9 shows that in 30 days the infected population of Kerala reached to 2100. Also we have observed real infected individual for 13 days overlap with our infected population curve. Figure 10 shows that the infected individuals in Kerala go to 50000 in 60 days from 21 March, 2020. But recent trends show that the infected individuals are far less than our expected number. This is because Kerala Government has taken proper strict preventing measures to stop spreading COVID-19 virus.

**Figure 9:**
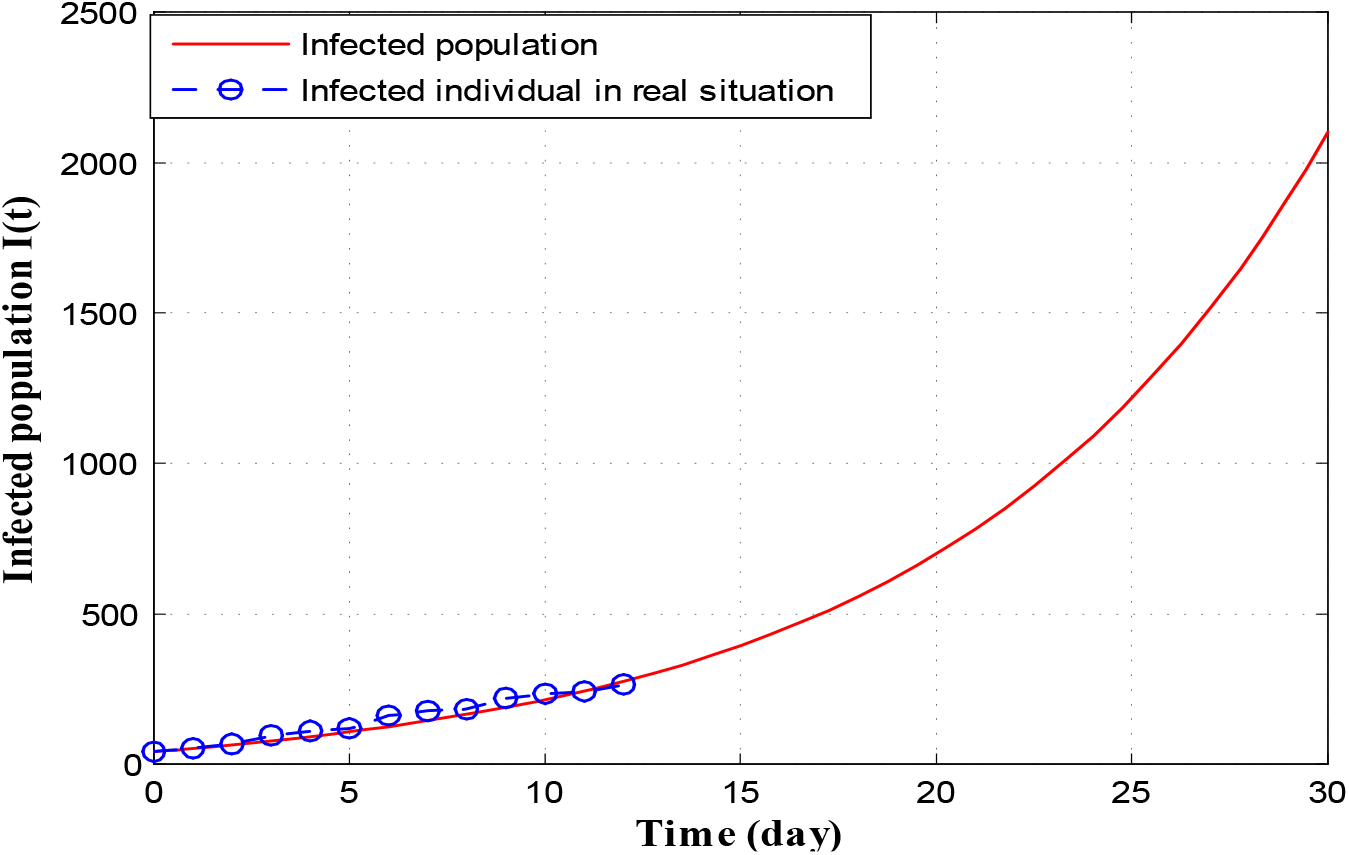
Time series plot of infected population in Kerala for *α =* 0.0000000 64 using data from Table 5 Table 6 and Table 7 for one month period from 22nd March, 2020.

**Figure 10:**
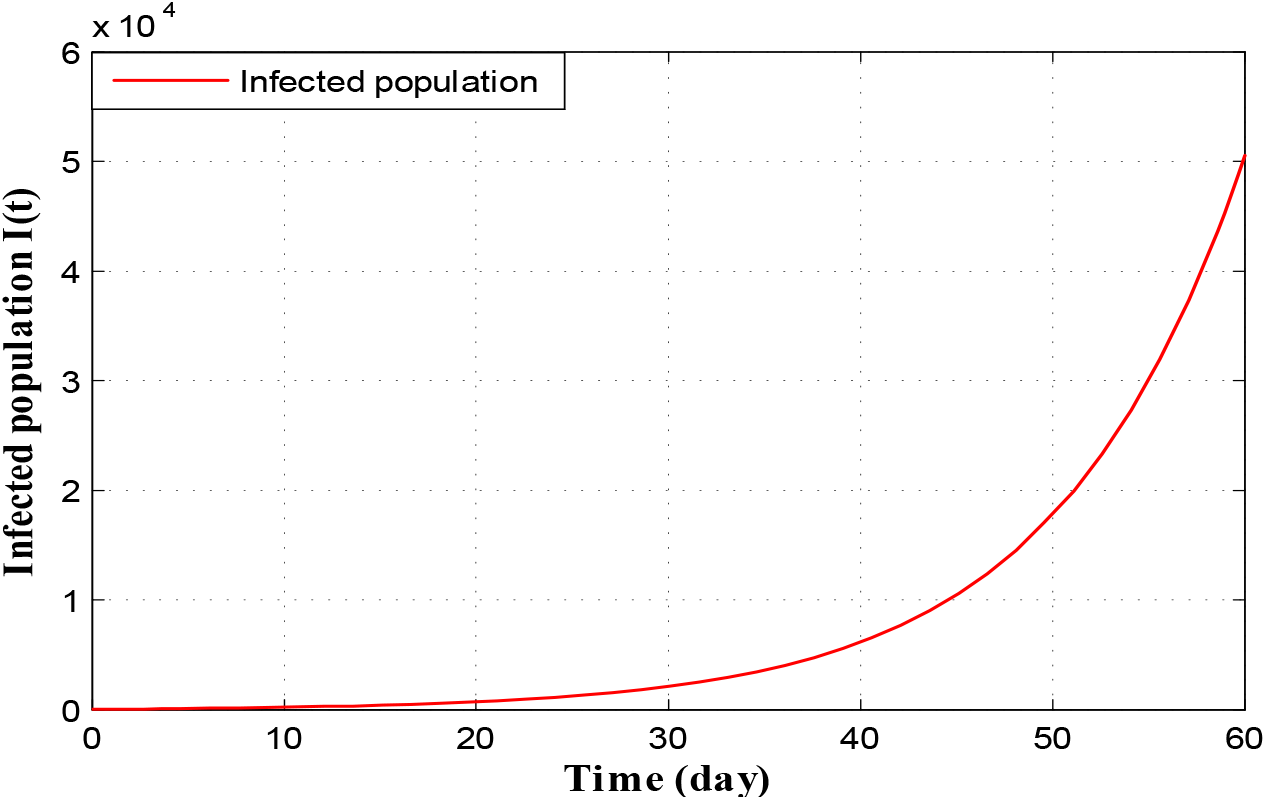
Time series plot of infected population in Kerala for *α =* 0.0000000 64 using data from Table 5 Table 6 and Table 7 for two months period from 22nd March, 2020

Following two figures gives the situation of COVID-19 in Uttar Pradesh

Figure 11 and Figure 12 depict that as per our construction infected population in 30 days and 60 days are 4400 and 620000 respectively starting from 21 March, 2020.

**Figure 11:**
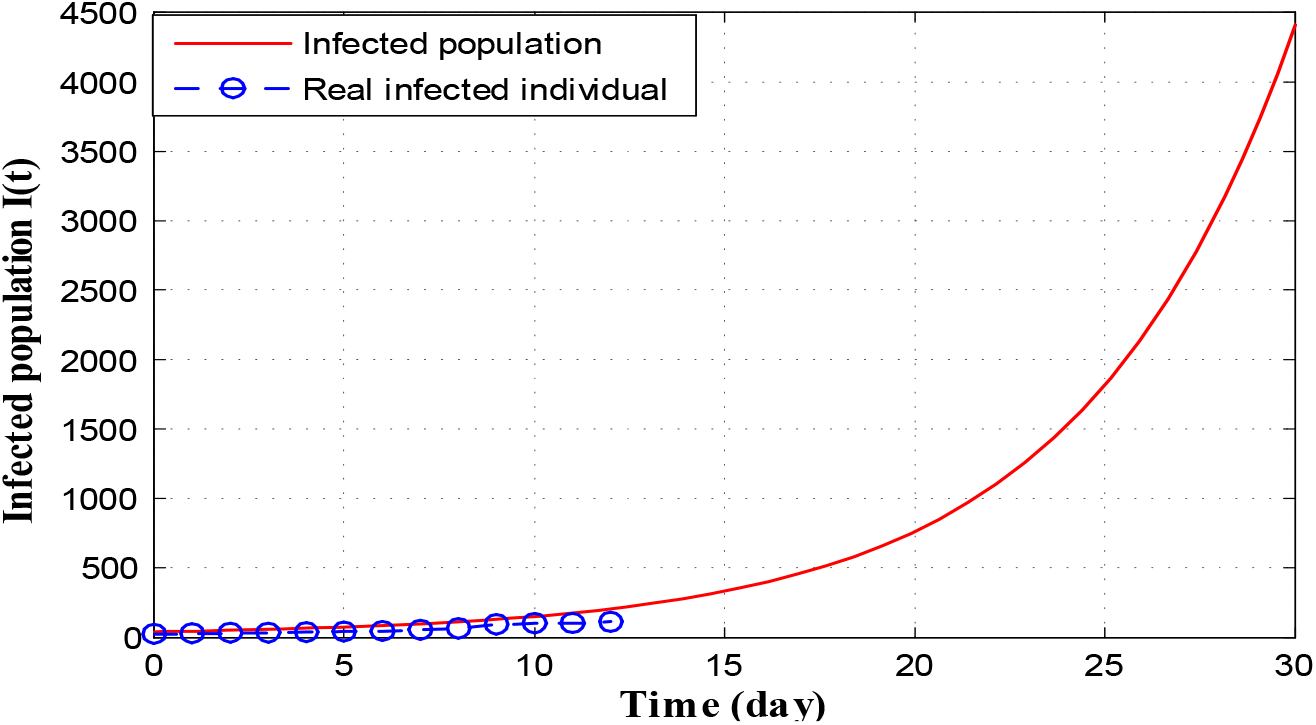
Time series plot of infected population in Uttar Pradesh for *α =* 0.000000020 using data from Table 5 Table 6 and Table 7 for one month period from 22nd March, 2020.

**Figure 12:**
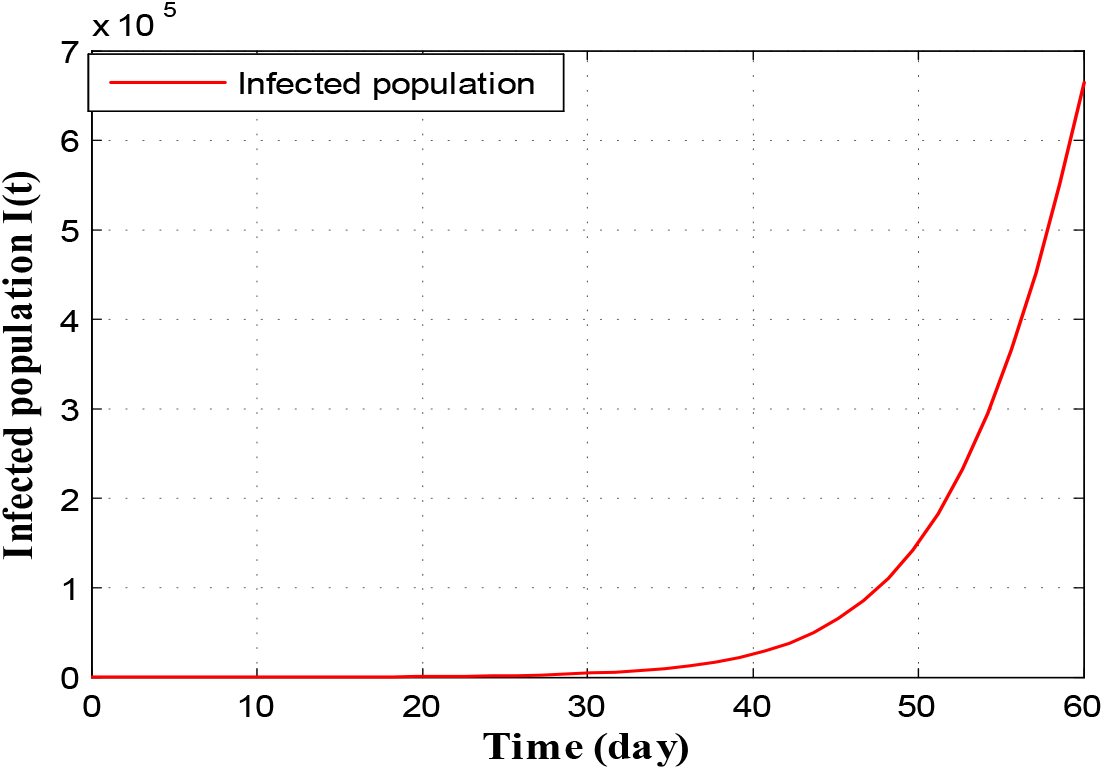
Time series plot of infected population in Uttar Pradesh for *α =* 0.000000020 using data from Table 5 Table 6 and Table 7 for two months period from 22nd March, 2020

Similarly, for the state Delhi we draw the following two figures

Figure 13 and Figure 14 depict that as per our construction infected population in Delhi during 30 days and 60 days are 4600 and 400000 respectively starting from 21 March, 2020.

**Figure 13:**
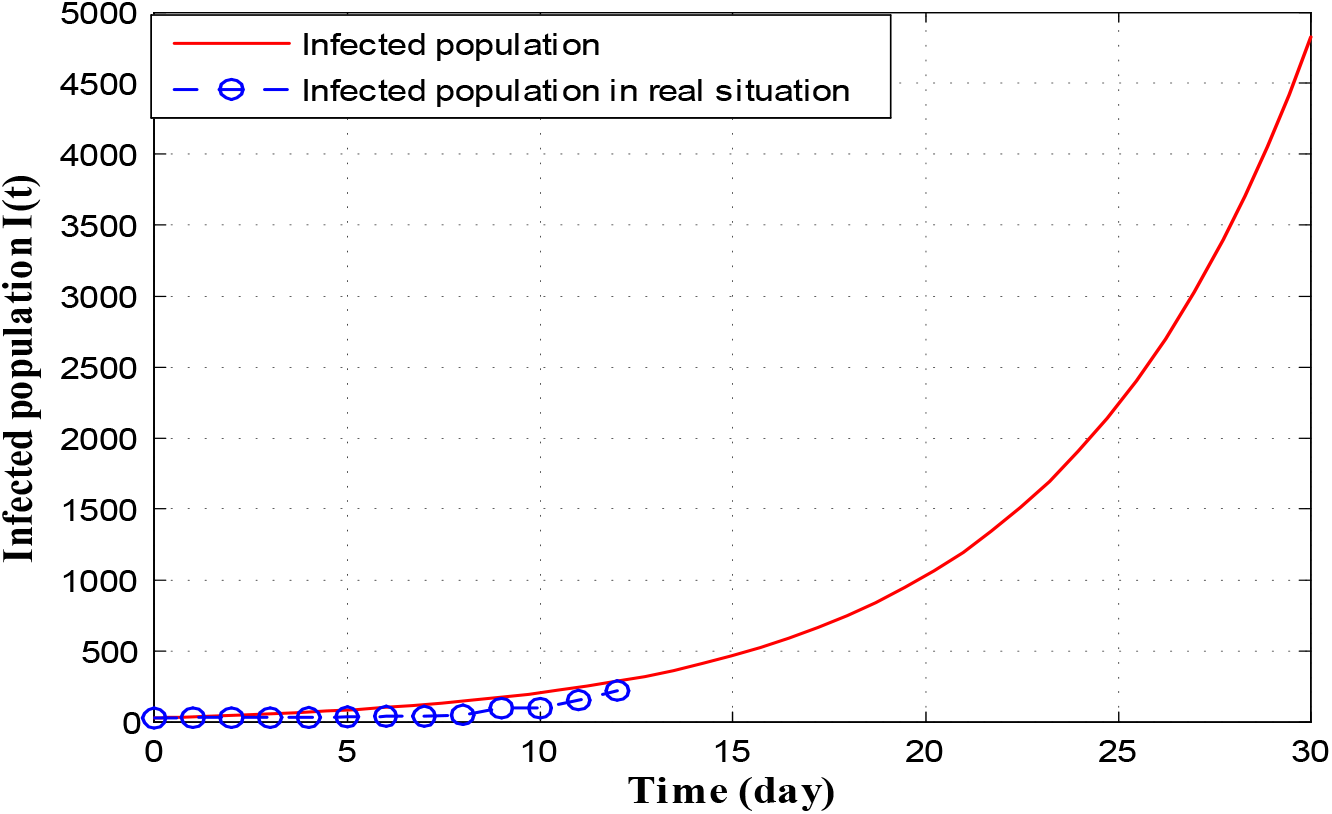
Time series plot of infected population in Delhi for *α =* 0.00000002 with initial conditions, parameter values are given in Table 5 Table 6 and Table 7 respectively for one month period from 21 March, 2020.

**Figure 14:**
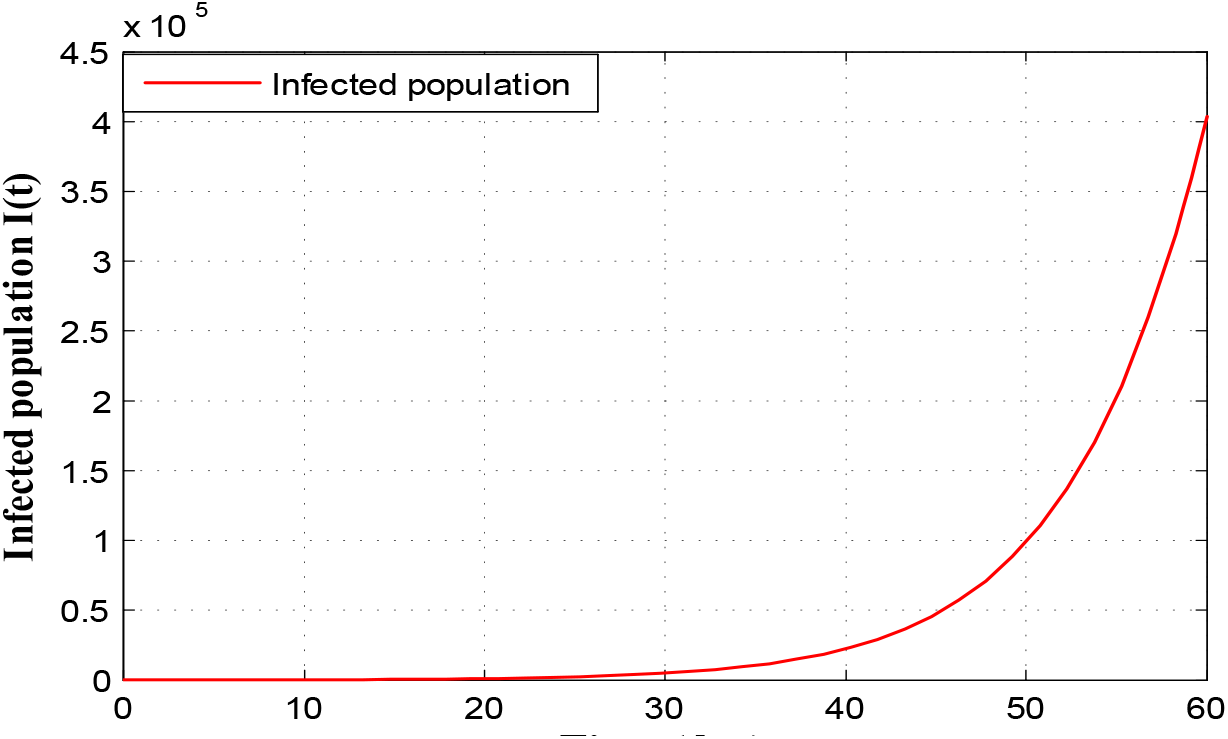
Time series plot of infected population in Delhi for *α =* 0.00000002 using data from Table 5 Table 6 and Table 7 for two month period from 21 March, 2020.

Finally COVID-19 pandemic situation in West Bengal can be expressed by the following two figures

As before Figure 15 and Figure 16 infer that as per our construction infected population in West Bengal during 30 days and 60 days are 1100 and 210000 respectively starting from 21 March, 2020.

**Figure 15:**
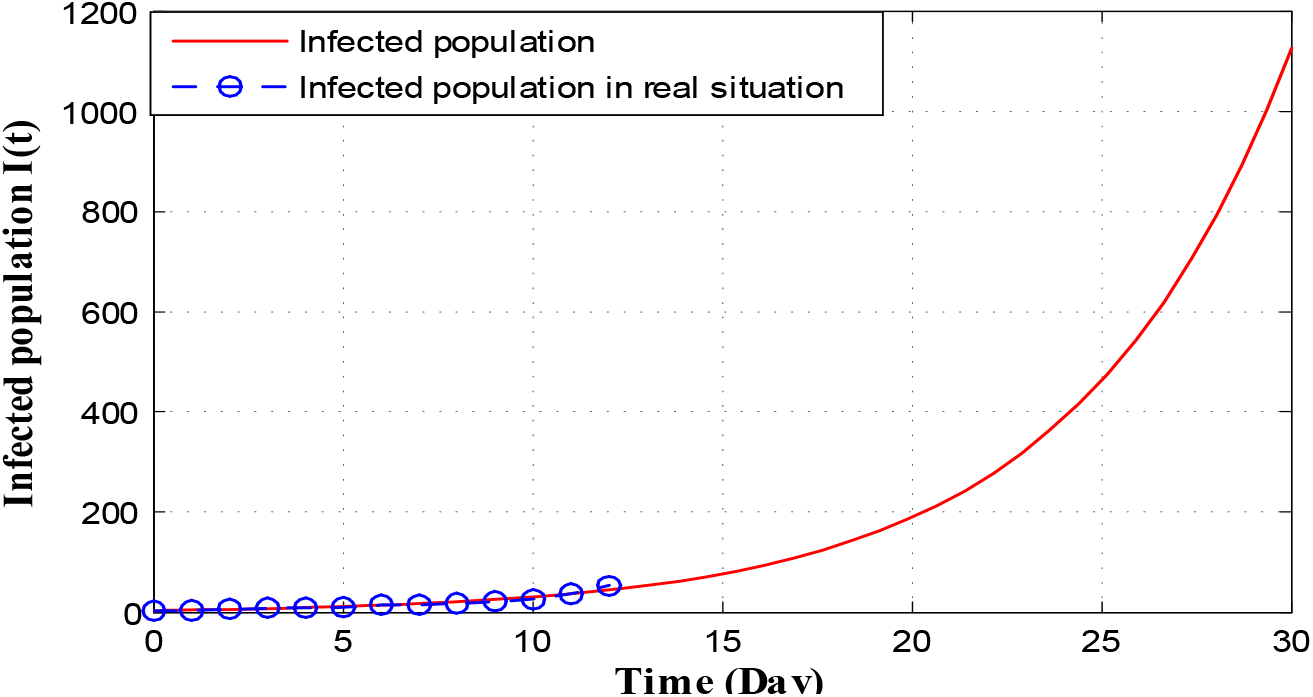
Time series plot of infected population in West Bengal for *α =* 0.0000000047 with initial conditions, parameter values are given in Table 5 Table 6 and Table 7 respectively for one month period from 21 March, 2020.

**Figure 16:**
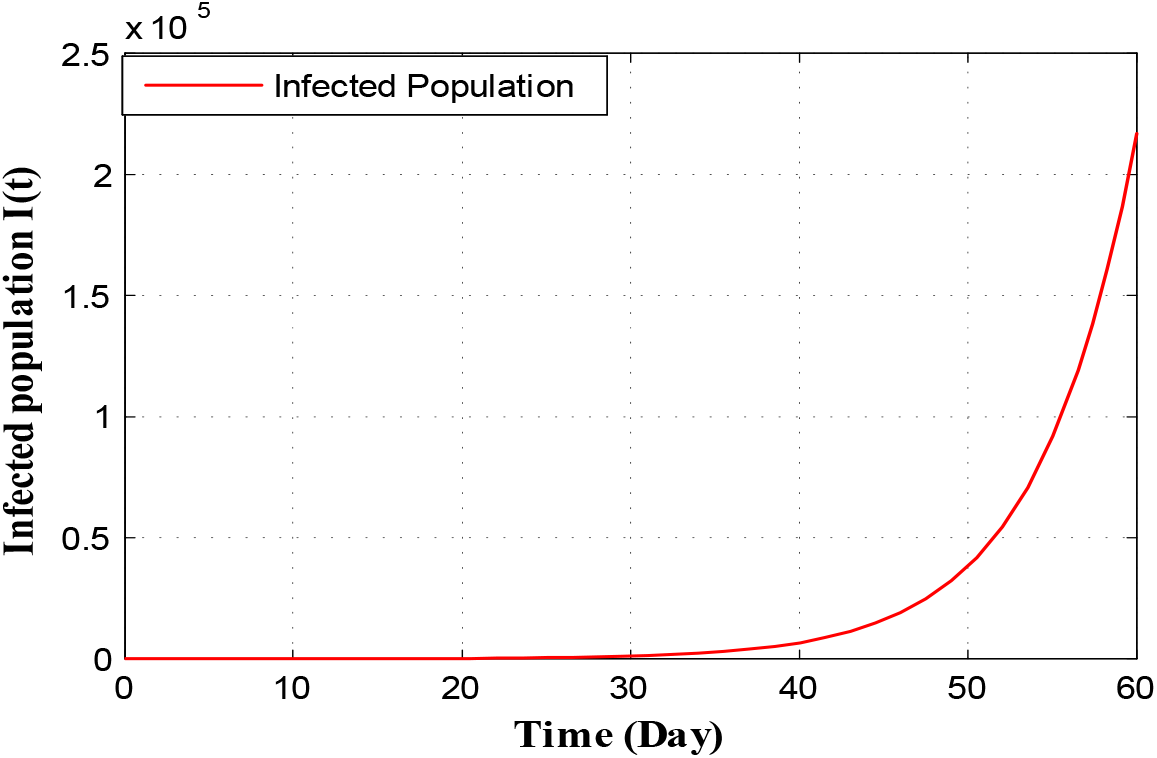
Time series plot of infected population in West Bengal for *α =* 0.0000000047 using data from Table 5 Table 6 and Table 7 for two months period from 21 March, 2020.

## 7 Conclusion and Discussion

In this current paper we have formulated and studied an epidemic model of COVID-19 virus which is transferred from human to human. So far the daily confirmed COVID-19 cases are increasing day by day worldwide. Therefore, prediction about infected individual is very much important for health concern arrangement of the citizens. It is also important to control spread rate of the COVID-19 virus with restricted supply. Our mathematical study is based on COVID-19 virus spread in India. We try to fit our model system to COVID-19 disease in India as per as limited data are available. The basic reproduction number *R*_0_ is calculated of our anticipated model. It is observed that when *R*_0_ < 1 then DFE *E*_0_ is globally asymptotically stable. Again, from sensitivity analysis of *R*_0_ we observe that the most sensitive parameter of our model structure is *α* (transmission rate from susceptible population to infected but not detected by testing population). Also the endemic equilibrium *E*_1_ exist and stable if *R*_0_ >1.

In this study our main aim is to predict mathematically the number of infected individual due COVID-19 virus in India. To fulfill our aim, we perform a detail numerical simulation of our proposed model in Section 6. We first simulate the COVID-19 situation in India, and thereafter situation of five different states (Maharashtra, Kerala, Uttar Pradesh, Delhi and West Bengal) are predicted numerically through graphical approach using Matlab software. We estimated the values of the parameter and the initial condition taken as per the information accessible on 21 March 2020.

We see that *α* is most sensitive parameter of our model system. So for different values of *α* infected population curve is presented in Figure 3. We observe that for *α =* 2.5 × 10^−l0^, the infected population curve is best fitted since it coincides with the actual infected individuals for 3 days starting from 2 March, 2020. Long term behavior of the infection is observed in figure 4. This figure shows us that as per the model construction as well as data available, the peak of infection is reached about 120 days after 21 march, 2020. Therefore, as per our prediction, the estimated date to reach the peak of infection is 21 July 2020 if the same restriction of measures is followed by the Indian Government as declared on 21 March 2020. Short term prediction about infected population is given via Figure 5 and Figure 6 respectively. Accordingly two of these figures predicted number of infected individual in India of COVID-19 patients in 30 and 60 days are 32000 and 3000000 respectively starting from 2 March 2020.

Now, we look upon the situation of different states. Figures 7 and 8 present that the number of infected population reaches to 4100 and 260000 (for *α =* 2.5 × 10^−9^) in next 30 and 60 days respectively starting from 21 March, 2020. For Kerala this number goes to 2100 and 50000 (for *α =* 0.0000000 64) respectively (see Figure 9 and Figure 10). In Uttar Pradesh, as per Figure 11 and Figure 12 (for *α =* 0.000000020) predicted numbers of infected individuals are 4400 and 620000. Total numbers of 4600 and 400000 people are infected in 30 and 60 days respectively in Delhi (look at Figure 13 and Figure 14). Finally, in West Bengal 1100 and 210000 people may be infected due to COVID -19 virus in 30 and 60 days respectively (see Figure 15 and Figure 16).

Therefore, analyzing all results of our proposed model and observing the situation of different countries, we may conclude that India may be in a big trouble in very near future due to COVID-19 virus. To avoid this big trouble, Indian Government should take stricter measures other than quarantine, lock down etc. So far the Indian Government are continuously changing its policy to protect India from COVID-19 virus. Recently, all districts of India are classified by Indian Government into three zones namely Red zones (hotspots), Orange zones (non hotspots) and Green zones (save zones). Preliminarily 170 districts of India are hotspots zones, where rapid testing facility is available for the public. As the time progress more strategies are applied by the Government of India as well as all state Governments to stop the spread of COVID-19 virus in India. Therefore, we may assume that if the Indian Government take proper step time to time then the infected number of population will be differ from our predicted number as time progress, and India will recover from this virus in recent future. Lastly, we say that public of India should help the Indian Government to fight against this dangerous COVID-19 as per the proposed model.

## Data Availability

Indian Council of Medical research
Ministry of Health and Family Welfare, Government of India 2020,

https://www.icmr.nic.in/.

https://www.mohfw.gov.in/

